# Saving costs and improving clinical outcomes: A two-year cost-utility analysis of emergency department care models for managing persons presenting with musculoskeletal pain using hybrid modelling

**DOI:** 10.1101/2025.09.05.25335189

**Authors:** Rose Gagnon, Naomi Hope Chouinard, Kadija Perreault, Simon LaRue, Simon Berthelot, Juliette Marchand, Komi Edem Gatovo, Luc J. Hébert, Jason R. Guertin

## Abstract

As access to primary healthcare remains challenging, numerous persons presenting musculoskeletal pain will visit the emergency department (ED) to receive care. Several approaches have been tried to optimize their management, such as the implementation of ED physiotherapy care models. However, no study has evaluated the efficiency of ED care models used to manage musculoskeletal pain beyond three months. This study evaluated the two-year efficiency of two ED care models (i.e., management by an emergency physician, management by a physiotherapist and an emergency physician) by conducting a cost-utility analysis using a hybrid mathematical model (decision tree + Markov model) from two different perspectives: Public Payer and Society. Data for this study came from a randomized clinical trial (n=78, #NCT04009369) and from the scientific and grey literature. A probabilistic approach was used to ensure the results’ robustness (Monte Carlo simulation, n=10,000 iterations). All costs were reported in CAD 2024 values. After two years, mean total cost per person for the physiotherapist and emergency physician management was lower than that of usual management by an emergency physician under both perspectives (Public Payer: $6,150 vs $6,840; Society: $30,978 vs $47,222). Mean quality of life gain was also higher in persons managed by a physiotherapist and an emergency physician (1.57 vs 1.47 quality-adjusted life years, QALYs). Management by a physiotherapist and an emergency physician was dominant under both perspectives. Integrating physiotherapists in EDs could result in long-term savings for the Public Payer and Society, while also helping to improve patients’ clinical outcomes.

## INTRODUCTION

Musculoskeletal (MSK) pain such as knee, neck and non-specific low back pain is very common [14]. In 2019 alone, more than 322,750,000 new persons presented MSK pain worldwide (incident cases) [31]. MSK pain is also a major cause of work absence and productivity loss [49]. As far back as the early 2000s, MSK pain already represented the second most important reason for consulting a doctor, and up to 20% of all primary care visits [49]. More recently, MSK pain has been found to account for more than 28% of all ambulatory medical visits [40]. Still, as access to primary care can be challenging in many countries, persons presenting with MSK pain also turn to the ED to receive care [43]. This limits continuity of care [29] and contributes to ED overcrowding [1].

Not surprisingly, costs associated with morbidity and disability related to MSK pain are enormous [49]. Chronic pain, including chronic MSK pain, is estimated to have direct and indirect costs ranging from $560 to $635 billion annually (2010 USD; $1,104 to $1,252 billion 2024 CAD), exceeding annual costs associated with heart disease, cancer and diabetes [26]. MSK pain alone is estimated to represent between 5.4% and 12.6% of a country’s total direct healthcare costs [34]. According to Park and al., chronic MSK pain could cost the United States’ healthcare system between $29,000 and $40,000 per patient per year (2016 USD; $51,900 to $71,600 2024 CAD) [39]. Still, the economic burden of chronic pain, including MSK pain, has been overlooked [44].

Knowing that persons presenting MSK pain often consult the ED, several approaches have been tried over the years to optimize their management. One of them is the integration of physiotherapists (PT) within the ED to manage these persons following nurse triage [5,9,15,24]. A growing number of studies report benefits associated with emerging PT care models in the ED [24,28,36]. Nonetheless, only two studies have examined the costs of these ED care models [35,42], and none have assessed their efficiency, that is, the costs related to the effectiveness of an intervention or care model. A recent study carried out by our research team found that, when compared to usual care by an emergency physician (EP) alone, combined management by a PT and an EP in the ED for persons consulting for MSK pain resulted in a greater gain in health-related quality of life at a lower cost, and was hence found to be dominant [21]. However, this study only looked at a three-month follow-up and thus cannot be used to assess longer-term efficiency.

Therefore, the aim of this study was to evaluate the efficiency over a two-year period of two ED care models for the management of persons presenting with musculoskeletal pain, namely management by an EP alone and management by a PT and an EP.

## METHODS

### Design

We performed a cost-utility analysis covering a two-year (24-month) period following an initial ED visit for MSK pain using a hybrid mathematical model comprising a decision tree and a Markov model [10,11]. The present study was part of a larger study on the efficiency of care models in the ED for MSK pain. Further details can be found in our published protocol [23].

Canada has a universal public healthcare system. However, not all health services are covered, including certain services that benefit persons presenting MSK pain (e.g., some outpatient physiotherapy services), which sometimes lead them to supplement their care in private healthcare settings. Hence, this cost-utility analysis was carried out using two different perspectives. On one hand, the Public Payer perspective included all costs paid by the public healthcare system (e.g., medical consultations, ED visits, hospitalizations). The Societal perspective, on the other hand, included all costs paid by the Public Payer, as well as costs not covered by the Public healthcare system that are related to insurance claims, personal out-of-pockets, productivity loss, and various complementary health-related governmental programs (e.g., public automobile insurance, workers’ compensation).

Our study was approved by the *CHU de Québec - Université Laval* Ethics committee (#MP-20-2019-4307). This manuscript complies with the *Consolidated Health Economic Evaluation Reporting Standards 2022* (CHEERS 2022) [27].

### Description of the Hybrid Mathematical Model

The hybrid mathematical model used to perform the cost-utility analysis combined a decision tree portion, focussing on the first three months, as well as a Markov model which covered the last 21 months. The choice of limiting our time horizon to 24 months was influenced by the paucity of scientific literature on the natural evolution of musculoskeletal pain, and by the short follow-up times of studies evaluating physiotherapy care models for managing persons presenting with musculoskeletal pain (six months to two years, e.g. [6,16]). More information on this choice can be found in the Supplementary material (Table S1).

The decision tree portion of the model (0 to 3 months) was based on observed data collected during the three-month follow-up of a pragmatic pilot randomized clinical trial (RCT) (n = 78, 18-80 years old, Canada, #NCT04009369) carried out in one of the EDs of the *CHU de Québec - Université Laval*. The aim of this pragmatic RCT was to compare the effects of care provided by a PT and an EP with that of care provided by an EP alone on clinical outcomes (e.g., pain, pain interference), healthcare resources used, health-related quality of life and healthcare costs. Management by an EP alone consisted of usual care, i.e., EP assessment following nurse triage, and intervention as deemed appropriate. The PT and EP care model, in turn, included initial management by a PT following nurse triage and subsequent management by an EP in order to comply with hospital bylaws in effect at the time of the study. The EP was free to take the PT’s management into account in their own management. In order to be recruited, persons presenting to the ED had to 1) present MSK pain (peripheral or vertebral), 2) be aged between 18 and 80 years old, and 3) be assigned a P3 (urgent), P4 (semi-urgent) or P5 (non-urgent) triage category according to the Canadian Triage and Acuity Scale [3]. At each follow-up post ED visit (1 and 3 months), participants were asked to complete a standardized health resource utilization questionnaire to compile all incurred costs, and questionnaires on clinical outcomes (e.g., Numeric Pain Ratin Scale [13], Brief Pain Inventory [47]). They were also asked to complete the EQ-5D-5L to detail any changes in their health-related quality of life [18]. More information on the RCT and its results can be found in Gagnon et al. [24].

The remaining 21 months of the hybrid model (Markov model portion) were modeled using parameters collected from the scientific and grey literature by RG, NHC and KEG, as well as from the previous RCT. All parameters collected from the literature and used in the constitution of the hybrid model were derived from studies with samples complying with the previous RCT’s inclusion criteria. Further details on the parameters and methodological choices that guided the constitution of the hybrid model are presented in Table S1 (Supplementary material). Both parts of the model are further described below.

#### Decision Tree

Figure 1A presents the decision tree used in the study. The decision tree’s transition probabilities were based on each participant’s pain level measured at both 1- and 3-month follow-ups during the RCT (Table S1, Supplementary material). The choice of discriminant threshold used to move participants through the decision tree was based on the minimum detectable change (MDC) of the Numeric Pain Rating Scale (scale used in the RCT to measure pain level), which is 2/10 [13]. All participants who reported a pain level of 3 or more out of 10 during the RCT follow-up were thus considered to present active MSK pain, while participants who reported a pain level of 0/10, 1/10 or 2/10 (i.e., below the scale’s MDC) were considered to be pain free.

**Figure 1.**
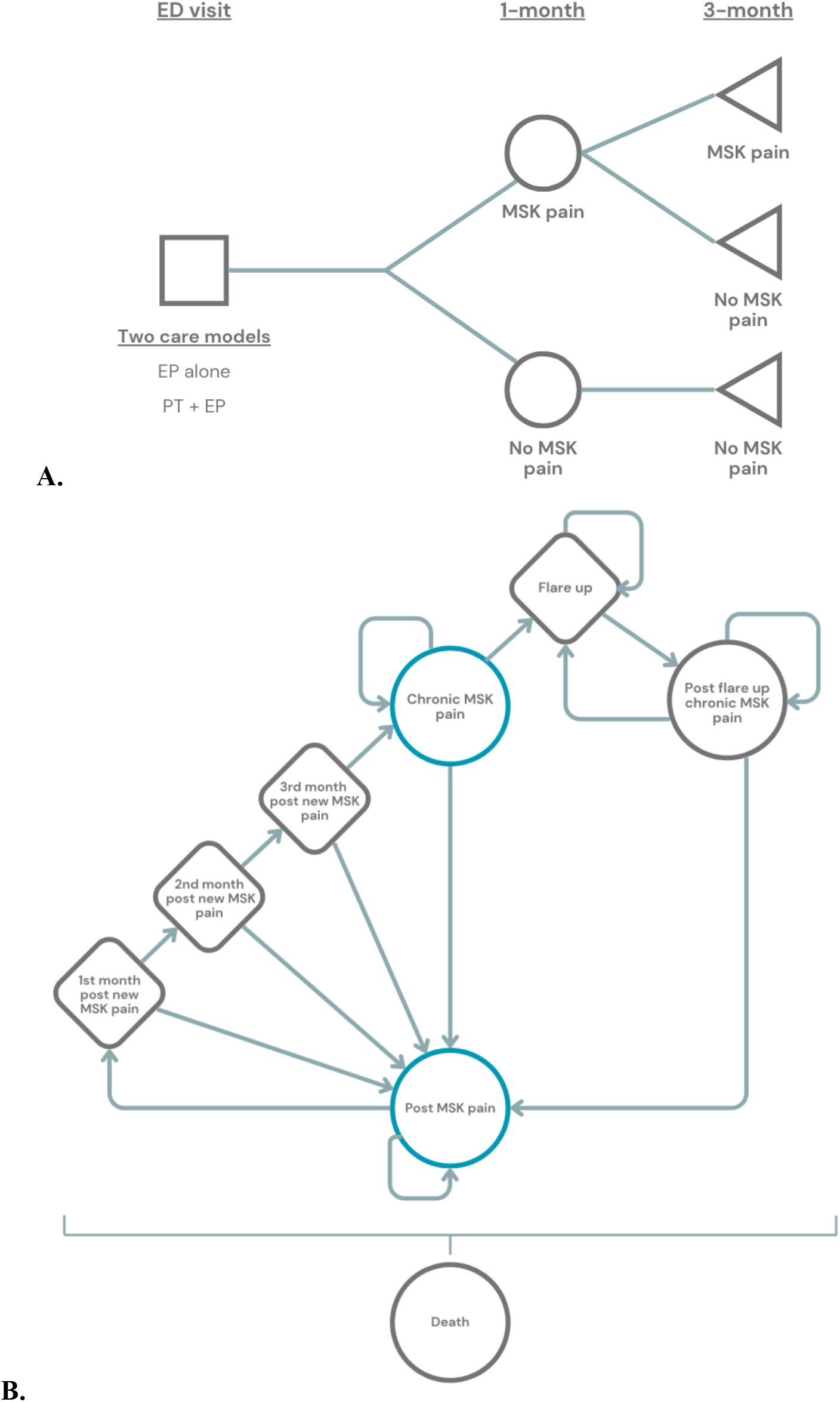
Hybrid mathematical model **A**.Decision tree *Circle: chance node, Triangle: terminal node* **B**.Markov model ED: emergency department, EP: emergency physician, PT: physiotherapist, MSK: musculoskeletal, 1^st^: first, 2^nd^: second, 3^rd^: third

Following the initial ED visit, all RCT participants with a pain level of 3/10 or more at the 1-month follow-up were considered as still presenting MSK pain and thus were considered to move to the “MSK pain” chance node at 1-month. Conversely, all participants who no longer presented pain moved to the “No MSK pain” chance node. The same rationale was applied at the 3-month follow-up for all RCT participants within the “MSK pain” chance node at 1-month. As no participant had developed new MSK pain between the 1- and 3-month follow-ups during the RCT, all participants who had moved to the “No MSK pain” node at 1-month moved to the “No MSK pain” node at 3-month.

These various steps were performed for each care model studied to derive 1) specific transition probabilities, and 2) specific conditional probabilities of being in a given terminal node (i.e., “MSK pain” or “No MSK pain”). The proportion of persons still presenting MSK pain after three months for each care model was then used to determine the number of persons in each state at the start of the Markov model. More information on the transition probabilities obtained and the distributions used to model them can be found in Figure S1 and Table S2 (Supplementary material).

Health-related quality of life data used for the decision tree were derived from values obtained within the previous RCT using the EQ-5D-5L. Scores were first transformed into utility scores using the Canadian algorithm [50]. The distribution of utility scores for each node was derived using mean utility scores of RCT participants who transitioned in each specific node. The total gain in quality-adjusted life years (QALY) per person over three months was then calculated according to their path in the decision tree using area-under-the-curve analyses [33].

Regarding costs, health resources used, as measured by the standardized RCT questionnaire, included new ED visits for the same condition, hospitalizations, consultations with doctors and/or health professionals, imaging, medication and walking aids/orthoses. Costing of these resources was carried out using scientific and grey literatures; more details can be found in our previous publications [21,25]. Mean cost per person was obtained by summing the mean node cost per node each RCT participant passed through in the decision tree.

#### Markov Model

The Markov model developed for this cost-utility analysis is shown in Figure 1B. It was designed to reflect as closely as possible the expected typical evolution of MSK pain over a period of four to 24 months. The model was validated by a panel of clinical experts not involved in its development during a formal elicitation process (90-minute meeting) to ensure its content validity (Table S1, Supplementary material).

Following their transition through the decision tree, RCT participants could begin their progression through the model in two different states: “Post MSK pain” and “Chronic MSK pain” (in blue, Figure 1B). Any participant who ended up in the two “No MSK pain” terminal nodes after three months in the decision tree began their passage through the Markov model in the “Post MSK pain” state. Conversely, all RCT participants who were identified as still having pain at the 3-month follow-up (i.e., “MSK pain” terminal node) started in the “Chronic MSK Pain” state. This methodological choice was based on international definitions of chronic pain as being pain lasting for at least three months (Table S1, Supplementary Material) [48].

RCT participants in the “1^st^”, “2^nd^” and “3^rd^ month post new MSK pain” states were forced to transit to a new state at each 1-month cycle (next state, return to “Post MSK pain” state or death). Indeed, these temporary states were added to better reflect the clinical evolution of musculoskeletal pain leading to the development of chronic musculoskeletal pain. Figure S2 presents the Markov model including all transition probabilities.

As for outcome data, mean utility scores and costs used for the “1^st^”, “2^nd^”, and “3^rd^ month post new MSK pain” states were derived from our RCT data. Mean utility scores and costs for all the other states were extracted from the scientific literature (Table S3, Supplementary material). Mean QALY gain per person and per cycle was obtained by multiplying the mean utility score specific to the state in which they were by the length of a cycle in years (one month: 1/12 year). In addition to all costs already considered in the decision tree, data from the literature allowed us to include costs associated with productivity loss (Societal perspective).

### Data Analyses

All costs and utility scores were analysed using descriptive statistics (means). These same costs and utility scores were used to derive an incremental cost-effectiveness ratio (ICER) per perspective to assess the cost-utility of the two care models considered. All costs are reported in 2024 Canadian dollars (2024 conversion rate, 1 USD = 1.3698 CAD, Bank of Canada). All costs incurred in the second year of the model were discounted at a rate of 1.5%, as suggested by current Canadian economic evaluation guidelines [8].

#### Uncertainty Evaluation

The hybrid mathematical model was created using a probabilistic approach to ensure the robustness of the results obtained (Monte Carlo Simulation, n = 10,000 iterations). Uncertainty around the ICERs is reported using cost-effectiveness planes and cost-effectiveness acceptability curves specific to each perspective. Besides the main analysis scenario, four other worst-case analysis scenarios were carried out, in which only the decision tree’s transition probabilities of persons managed by a PT and an EP were modified (i.e., increase in their probability of presenting persistent MSK pain), to further test the results’ robustness.

More information on the parameters used, the rationale behind their selection and the types of distributions used in the probabilistic sensibility analyses can be found in Tables S2 and S3 (Supplementary material). In addition, Table S4 details the different cost types included in each perspective. More details on the methods used to characterize the uncertainty associated with the analyses performed are presented in Table S5. All analyses were performed using R statistical software (version 4.4.2, R Foundation).

## RESULTS

### Characteristics of Participants Recruited During the Randomized Clinical Trial

Table 1 presents the baseline sociodemographic characteristics of participants recruited for the RCT. Characteristics were very similar between the two groups. However, participants managed by a PT and an EP were significantly younger and more likely to be a female (Table 1). The cost and utility distributions used to develop the model were therefore based on age- and sex-adjusted costs and utility scores.

**Table 1.** Baseline sociodemographic characteristics of the randomized clinical trial participants (n=78)

| Characteristics | Usual care<br>EP alone | New care model<br>PT + EP |
| --- | --- | --- |
| Number of participants, *n* (%) | 38 (48.7) | 40 (51.3) |
| Age (yr), mean (SD) | 44.1 (17.3) | 36.6 (17.3) |
| Sex, *n* females (%) | 12 (31.6) | 22 (55.0) |
| Other health condition, *yes* (%) | 23 (60.5) | 26 (65.0) |
| Localisation of MSK pain, *n* (%) |  |  |
| Upper/lower limb | 19 (50.0) | 23 (57.5) |
| Spine | 19 (50.0) | 17 (42.5) |
| Weeks since onset, *n* (%) |  |  |
| 0 to 2 | 32 (84.2) | 30 (75.0) |
| 3 to 12 | 1 (2.6) | 4 (10.0) |
| More than 12 | 4 (10.5) | 4 (10.0) |
| Don't know | 1 (2.6) | 1 (2.5) |
| Family physician, *yes* (%) | 30 (78.9) | 33 (82.5) |
| Pain level <sup>a</sup>, /10 (SD) | 6.7 (2.2) | 6.9 (2.0) |
| Pain interference <sup>b</sup>, /10 (SD) | 4.4 (1.8) | 4.1 (2.3) |
| Pain catastrophizing <sup>c</sup>, /100 (SD) | 22.4 (11.8) | 18.3 (13.2) |

EP: emergency physician, PT: physiotherapist, yr: year, SD: standard deviation, MSK: musculoskeletal
<sup>a</sup> Measured using the Numeric Pain Rating Scale [4,13]
<sup>b</sup> Measured using the Brief Pain Inventory [37,47]
<sup>c</sup> Measured using the Pain Catastrophizing Scale – Canadian French [19]

### Clinical Evolution

Proportion of persons presenting persistent MSK pain after three months was 7.4% for the PT and EP care model, and 31.4% for the usual care model (Figure S1, decision tree). After two years in the model, 35.9% of persons managed via the EP alone care model were in the “Post flare up chronic MSK pain” state, compared to 7.2% of persons managed via the PT and EP care model.

### Health-related Quality of Life

After three months (decision tree portion), persons managed by the PT and EP had a lower QALY gain per person than those managed in the usual way by an EP alone (Differential: −0.0005, Table 2). However, after two years, the group managed via the PT and EP care model showed a higher QALY gain per person than the EP group (1.5676 vs. 1.4657, Table 2).

**Table 2.** Average total cost and QALY gain per person, per intervention and per perspective

|  | Mean cost <sup>a</sup> (95% CI) |  |  |  |  |  |
| --- | --- | --- | --- | --- | --- | --- |
|  | Public Payer perspective |  |  | Societal perspective |  |  |
|  | Usual care<br>EP alone | New care model<br>EP + PT | Differential | Usual care<br>EP alone | New care model<br>EP + PT | Differential |
| **Average total costs** |  |  |  |  |  |  |
| 3-month | 1,079<br>(461; 2,205) | 716<br>(345; 1,320) | -363<br>(-1,111; 25) | 1,841<br>(874; 3,522) | 1,223<br>(625; 2,185) | -619<br>(-1,766; 5) |
| 2-year | 6,840<br>(5,097; 10,208) | 6,150<br>(4,775; 8,541) | -690<br>(-3,030; 155) | 47,222<br>(27,638; 98,590) | 30,978<br>(22,193; 49,086) | -16,244<br>(-56,410; -1,970) |
| **Average total QALY gain** |  |  |  |  |  |  |
| 3-month | 0.2057<br>(0.1590; 0.2482) | 0.2052<br>(0.1698; 0.2342) | -0.0005<br>(-0.0300; 0.0304) | 0.2057<br>(0.1590; 0.2482) | 0.2052<br>(0.1698; 0.2342) | -0.0005<br>(-0.0300; 0.0304) |
| 2-year | 1.4657<br>(1.0041; 1.7790) | 1.5676<br>(1.0920; 1.8460) | 0.1019<br>(-0.0780; 0.3489) | 1.4657<br>(1.0041; 1.7790) | 1.5676<br>(0.9509; 1.8629) | 0.1019<br>(-0.0780; 0.3489) |
| **2-year ICER** | **- 6,770 / QALY [Dominant]** |  |  | **- 159,418 / QALY [Dominant]** |  |  |

CI: confidence interval, EP: emergency physician, PT: physiotherapist, QALY: quality-adjusted life years, ICER: incremental cost-effectiveness ratio
<sup>a</sup> All costs are in 2024 Canadian dollars

### Costs

#### Public Payer perspective

Mean total cost per person after three months was $716 for the PT and EP care model, and $1,079 for the usual care model. After two years, mean total cost per person increased to $6,150 and $6,840 for the PT + EP and EP alone groups respectively, for a difference in mean total cost per person between the two groups of -$690 per person (95% confidence interval (CI): -$3,030; $155).

#### Societal perspective

As with the Public Payer perspective, mean total cost per person after three months under the Societal perspective was lower for the PT and EP care model than for the usual care model ($1,223 vs. $1,841, differential (95%CI): -$619 (-$1,766; $5)). After two years, persons managed by a PT and an EP had an average societal cost of $30,978 per person, compared with $47,222 per person for those managed by an EP alone (Differential (95%CI): -$16,244 (-$56,410; -$1,970)).

### ICER

Management by a PT and an EP was found to be dominant under both perspectives, with ICERs of - $6,770 per QALY gained for the Public Payer perspective and -$159,418 per QALY gained for the Societal perspective. Figure 2 displays the cost-effectiveness planes from both perspectives. In both cases, management by a PT and an EP was dominant (i.e., more effective, less costly) in more than 80% of iterations. Cost-effectiveness acceptability curves obtained from the two perspectives are presented in Figure 3. For a willingness-to-pay of $0, management by a PT and an EP was the most cost-effective care model in almost 100% of iterations. This proportion of cost-effective iterations remained close to 100% for all willingness-to-pay thresholds tested within the Societal perspective, while it dropped to around 90% for a willingness-to-pay of $100,000 under the Public Payer perspective.

**Figure 2.**
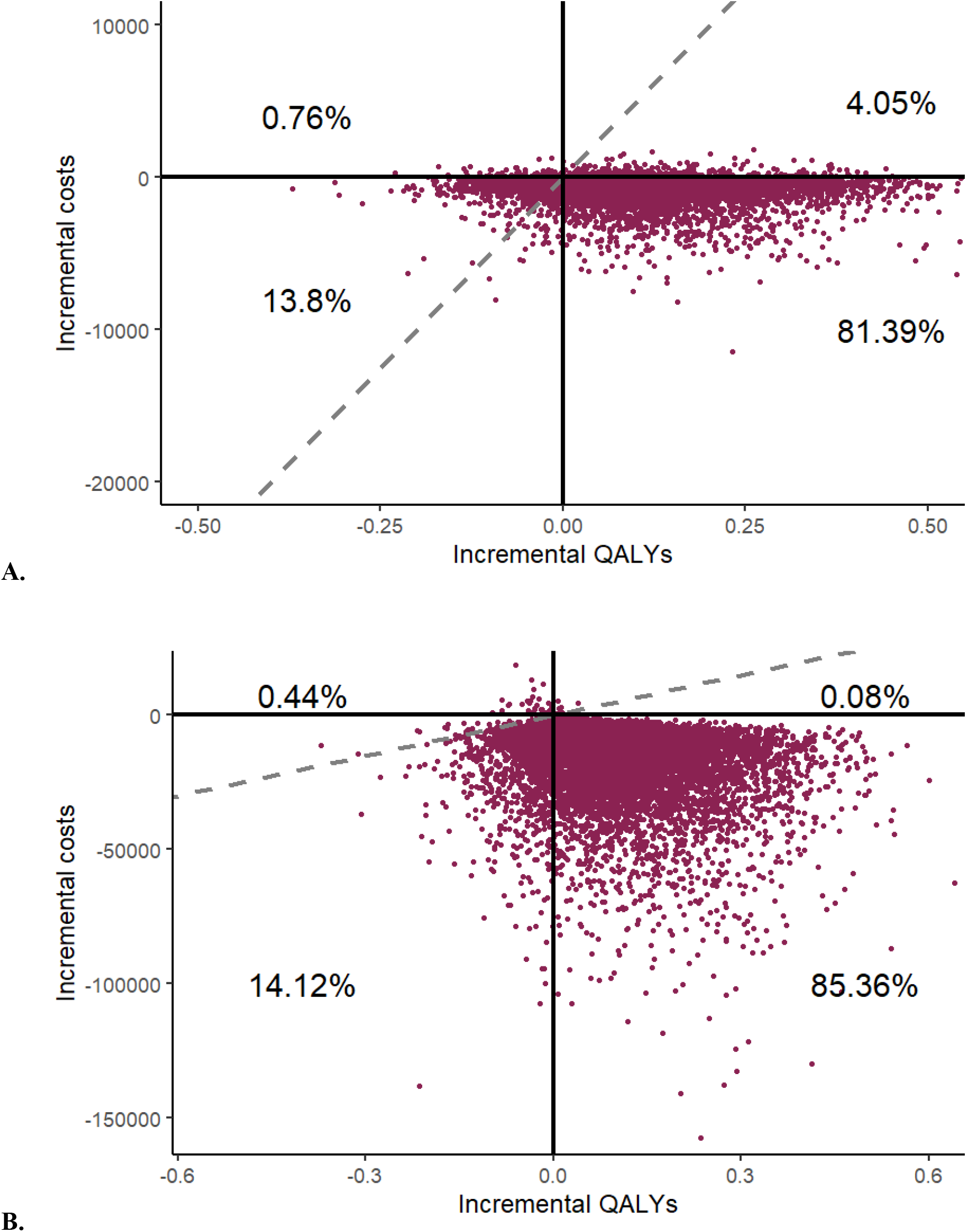
Cost-effectiveness plane for each perspective **A**.Public Payer perspective **B**.Societal perspective QALY: quality-adjusted life years

**Figure 3.**
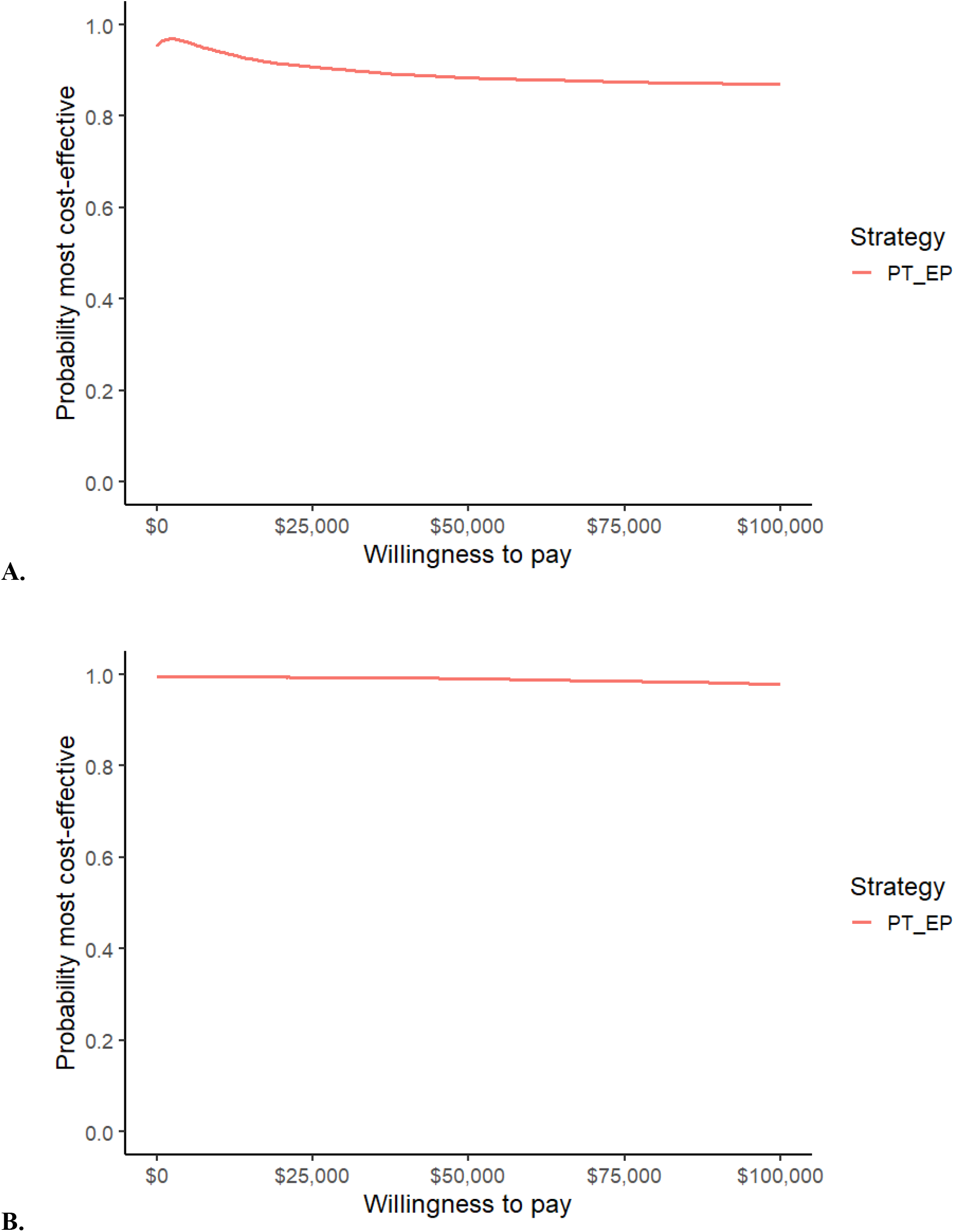
Cost-effectiveness acceptability curve for each perspective **A**.Public Payer perspective **B**.Societal perspective EP: emergency physician; PT: physiotherapist

#### Uncertainty

In three of the four worst-case scenarios tested, management by a PT and an EP was found to be dominant, with the percentage of dominant iterations under both perspectives ranging from 47% to 77% (Figures S3 to S5). However, in the most pessimistic scenario tested where the probability of having MSK pain at 1-month was doubled and the probability of having MSK pain at 3-month was 100% for all persons managed via the PT and EP care model (Fourth assumption, Figure S6), usual care by an EP dominated care by a PT and an EP in 44% and 73% of iterations when assessed from the Public Payer’s perspective and from the Societal perspective, respectively.

## DISCUSSION

The aim of this study was to evaluate, over a two-year horizon, the efficiency of two ED care models for the management of persons presenting with MSK pain. In our base case scenario, the care model consisting of management by a PT and an EP was found to be dominant when compared to management by an EP alone. All the worst-case scenarios analysed were also consistent with these results, except for the most pessimistic of the four (Fourth assumption, Figure S6; possibly less realistic), increasing the overall robustness of our conclusions.

The efficiency of the PT and EP care model may be largely explained by the fact that, after three months, only 7.4% of persons managed by a PT and an EP entered the Markov model in a state of chronic MSK pain, compared with 31.4% of those managed by an EP alone (Figure S1). Since the annual recovery rate for chronic pain used in the Markov model was estimated at 5.4% (monthly recovery rate: 0.46%) [17], many of those who entered the model in a state of chronic MSK pain remained in that state for the entire time horizon, which may have contributed significantly to the observed costs. In addition, many persons with chronic MSK pain will be at risk of experiencing a flare up of their condition (11.21% per month) [46], which is likely to have further increased the observed mean total cost per person. A broader implementation of care models such as management by a PT and an EP could potentially help reduce the incidence of chronic MSK pain, and the magnitude of its societal consequences, such as lost productivity, absenteeism and significant use of healthcare resources [12,20,30,51].

As previously mentioned, to our knowledge, no cost-effectiveness or cost-utility analysis of management by a PT and an EP in the ED has been carried out over a time horizon of more than three months. However, in primary care, a previous cost-utility analysis (Sweden) suggests that direct access PT management would be dominant when compared to usual management by a family physician over a one-year time horizon [6]. According to Bornhoft et al., PT management could help save the Public Healthcare System $30 per patient and Society $3,600 per patient (2017 EUR; $57 and $6,875 2024 CAD), in addition to improving health-related quality of life [6]. Some retrospective studies based on administrative data also showed that early PT management is associated with a reduction in total MSK pain-related costs (between $2,182 and $8,787 per patient) two years after initial management [12,20,32]. The results of these studies support the importance of further evaluating the impact of integrating different healthcare professionals in the ED, such as PTs, on healthcare costs and clinical outcomes.

MSK pain is a condition with a significant societal impact [7]. Based on a 2015 study, considering only the costs arising from lost productivity, the societal costs of MSK pain could be as high as 2% of the European gross domestic product [2]. Nonetheless, very few studies have examined the epidemiology and natural evolution of MSK pain [41]. Moreover, very few longitudinal studies have evaluated the impacts of care models put in place to manage persons presenting with MSK pain on the associated burden [22]. Although the present hybrid mathematical model is a promising step, the time horizon considered remains short, which is inconsistent with health economics guidelines recommending that a chronic disease be modeled over a lifetime horizon [8]. It is therefore possible that the current results underestimate the longer-term impact of the care models evaluated, given that the presence of chronic MSK pain increases the risk of developing other chronic diseases [7]. Considering that the burden of MSK pain is expected to continue increasing over the coming years due to an aging population, increasing obesity and physical inactivity [14,45], more high-quality longitudinal data will be needed to adequately plan health services for these persons.

This study has several limitations. The decision-tree portion of the hybrid mathematical model was based on data from a pilot pragmatic RCT with a small sample size (n=78) [24]. The small number of observations for certain cost types thus sometimes led to significant variability within the cost distributions. In addition, as discussed above, there is little literature reporting longitudinal data on costs and clinical outcomes of persons presenting with MSK pain. This had the effect of limiting 1) the inclusion of the same cost types within each state of the Markov model, 2) the time horizon considered and 3) the possibility of realizing stratified analyses (e.g., sex, age, type of MSK pain). Finally, some of the data included in the conception of the Markov model only included adults aged between 20 and 55 given the paucity of literature examining death rates among persons presenting musculoskeletal pain [38].

Despite these limitations, this study also has significant strengths. Part of the hybrid mathematical model was based on observed data rather than on data from the scientific literature, thereby increasing its validity. The Markov model was also conceptualized and validated by healthcare professionals with a good knowledge of the condition of interest. Where possible, several types of costs were included, such as those related to lost productivity and time, to better capture the true burden associated with MSK pain. Finally, although the data used showed considerable variability, several worst-case scenarios were performed to ensure the robustness of the results obtained.

## CONCLUSION

Results of this study indicate that, when compared to usual management by an EP alone, management by a PT and an EP would allow the Public Payer and Society to save money over a two-year period, while improving the health-related quality of life of persons presenting with MSK pain. Given the expected increase in the prevalence of MSK pain, the integration of such models of care in the ED could help decision-makers to optimize the use of their limited healthcare resources (e.g., human, financial) when planning health services for persons presenting with MSK pain.

## Data Availability

All data produced in the present study are available upon reasonable request to the authors.

