## Supplementary Material for "Saving costs and improving clinical outcomes: A two-year cost-utility analysis of emergency department care models for managing persons presenting with musculoskeletal pain using hybrid modelling"

**Table S1** Methodological choices made during the conception of the hybrid mathematical model

| Item | Methodological choice | Sources |
|---|---|---|
| Time horizon | The choice of the length of the time horizon for the hybrid mathematical model was based on a number of considerations:<br>1. To be considered chronic, musculoskeletal pain must last at least three months;<br>2. Approximately 30% of persons presenting musculoskeletal pain will report pain and functional disability for more than 12 months after the onset of their pain;<br>3. Very few studies on the epidemiology of musculoskeletal pain (e.g., long-term prevalence and incidence, expected natural evolution, long-term impact on the development of other health conditions, etc.) have been carried out to date;<br>4. The few clinical studies evaluating the impact of physiotherapy management had follow-up durations ranging from six to 24 months. | Kovacevic, Kogler et al.;[1]<br>Caffrey, Smart et al.;[2]<br>Downie, McRitchie et al.;[3]<br>Lau, Chow et al.;[4]<br>Bornhöft, Larsson et al.[5] |
| Cycle length | The cycle length initially envisaged in the research protocol was two weeks, in order to better capture the sometimes rapid evolution of musculoskeletal pain in the first few weeks. However, the limited epidemiological literature available did not allow for a cycle shorter than one month. | Gagnon, Hébert et al.[6] |
| Selection of outcomes | As per the original research protocol, the different states of the Markov model were intended to be based on the level of pain interference with function, and not on changes in pain status during follow-ups. However, in addition to being relatively scarce, the literature on the natural evolution of musculoskeletal disorders/pain does not use a consensual definition to define disability and function. Thus, the definitions and measurement scales used varied from study to study, making it difficult to create a model with a uniform definition. It was therefore decided to base the model on the definitions of acute, subacute and chronic pain, which are internationally standardized and applied more uniformly across the different epidemiological studies. | Gagnon, Hébert et al.[6]<br>Treede, Rief et al.[7] |
| Valuation of outcomes | The various questionnaires and scales used to measure the variables of interest were previously selected as part of a pilot pragmatic randomized clinical trial from which some of the data required for the present study were extracted. | Gagnon, Perreault et al.[8] |
| Model validation | Validation of the Markov model was carried out with three experts in the condition of interest (musculoskeletal pain) during a 90-minute meeting. During this meeting, the experts received basic training on the methodological principles of Markov models and were then asked to comment on the content validity of the different states and transition probabilities used. | |
| Approaches to engage clinicians and decision makers | The pilot pragmatic randomized clinical trial from which some of the necessary data for the hybrid mathematical model is drawn was co-developed in collaboration with managers and clinicians from the host setting (*CHU de Québec - Université Laval*). | Gagnon, Perreault et al.[8] |

**Table S2** Parameters used to perform the cost-utility analysis – Decision tree

| Parameter | Values | 95% CI | Probability Density Function |
|---|---|---|---|
| **Transition probabilities** | **%** | | |
| **1-month** | | | |
| ***EP alone*** | | | |
| Probability of having MSK pain | 62.4 | 45.2-78.1 | Beta |
| Probability of having no MSK pain | 37.6 | 21.9-54.8 | |
| ***PT + EP*** | | | |
| Probability of having MSK pain | 25.8 | 12.4-42.5 | Beta |
| Probability of having no MSK pain | 74.2 | 57.5-87.6 | |
| **3-month** | | | |
| ***EP alone*** | | | |
| Probability of having MSK pain when having pain at 1-month | 50.3 | 26.5-73.2 | Beta |
| Probability of having no MSK pain when having pain at 1-month | 49.7 | 26.8-73.4 | |
| Probability of having no MSK pain at 3-month when having no MSK pain at 1-month | 100.0 | --- | |
| ***PT + EP*** | | | |
| Probability of having MSK pain when having pain at 1-month | 28.5 | 4.3-63.9 | Beta |
| Probability of having no MSK pain when having pain at 1-month | 71.5 | 36.1-95.7 | |
| Probability of having no MSK pain at 3-month when having no MSK pain at 1-month | 100.0 | --- | |
| **Costs, per node** | **$ [a]** | | |
| **1-month** | | | |
| ***Canadian Public Payer*** | | | |
| MSK pain | 625.05 | 177.95-1,471.09 | Gamma |
| No MSK pain | 434.26 | 160.17-891.43 | |
| ***Canadian Society*** | | | |
| MSK pain | 988.99 | 326.45-2,180.09 | Gamma |
| No MSK pain | 537.25 | 216.42-1,046.41 | |
| **3-month** | | | |
| ***Canadian Public Payer*** | | | |
| MSK pain (1-month: MSK pain) | 158.22 | 58.65-329.56 | Gamma |
| No MSK pain (1-month: MSK pain) | 210.20 | 21.54-932.43 | |
| No MSK pain (1-month: No MSK pain) | 20.91 | 6.50-45.51 | |
| ***Canadian Society*** | | | |
| MSK pain (1-month: MSK pain) | 279.04 | 96.23-573.96 | Gamma |
| No MSK pain (1-month: MSK pain) | 774.75 | 140.72-2,187.73 | |
| No MSK pain (1-month: No MSK pain) | 203.35 | 15.74-787.77 | |
| **Effectiveness, per node** | **Mean utility score** | | |
| **1-month** | | | |
| MSK pain | 0.647 | 0.294-0.917 | Beta |
| No MSK pain | 0.834 | 0.537-0.988 | |
| **3-month** | | | |
| MSK pain (1-month: MSK pain) | 0.707 | 0.456-0.901 | Beta |
| No MSK pain (1-month: MSK pain) | 0.717 | 0.416-0.936 | |
| No MSK pain (1-month: No MSK pain) | 0.908 | 0.801-0.975 | |

CI: confidence interval; %: percentage; EP: emergency physician; MSK: musculoskeletal; PT: physiotherapist; $: dollars
[a] All monthly costs are in 2024 Canadian dollars

**Table S3** Parameters used to perform the cost-utility analysis – Markov model

| Parameter | Values | 95% CI | Probability Density Function | Rationale | Source |
|---|---|---|---|---|---|
| **Transition probabilities** [a] | | % | | | |
| ***Post initial MSK pain*** | | | | | |
| Probability of having no other MSK pain | 96.71 | 92.44-99.22 | Dirichlet | Considering that all transition probabilities for each state must add up to 100%, this probability was obtained by subtracting this state's other transition probabilities from 100%. | Not applicable |
| Probability of having new MSK pain | 3.28 | 0.77-7.54 | | In this study, 33% of participants had a recurrence of their low back pain within a year. | Machado, Maher et al.[9] |
| Probability of dying while having no MSK pain | 0.011 | 0-0.061 | | Probability of death from any cause in the general Canadian population aged 20-55 years. This age range was chosen to align with the all-cause death rates for persons presenting chronic pain available in the literature. | Statistics Canada[10] |
| ***First month post new MSK pain*** | | | | | |
| Probability of MSK pain resolving after one month | 24.99 | 16.98-33.74 | Dirichlet | Considering that all transition probabilities for each state must add up to 100%, this probability was obtained by subtracting this state's other transition probabilities from 100%. | Not applicable |
| Probability of MSK pain lasting more than one month | 75.00 | 66.25-83.01 | | Proportion of study participants whose MSK pain persisted after one month. | Heneweer, Aufdemkampe et al.[11] |
| Probability of dying while presenting new MSK pain | 0.011 | 0-0.061 | | Probability of death from any cause in the general Canadian population aged 20-55 years. This age range was chosen to align with the all-cause death rates for persons presenting chronic pain available in the literature. | Statistics Canada[10] |
| ***Second month post new MSK pain*** | | | | | |
| Probability of MSK pain resolving after two months | 46.39 | 36.65-56.26 | Dirichlet | Considering that all transition probabilities for each state must add up to 100%, this probability was obtained by subtracting this state's other transition probabilities from 100%. | Not applicable |
| Probability of MSK pain lasting more than two months | 53.60 | 43.71-63.35 | | Proportion of study participants whose MSK pain persisted after two months. | Heneweer, Aufdemkampe et al.[11] |
| Probability of dying while presenting new MSK pain | 0.011 | 0-0.061 | | Probability of death from any cause in the general Canadian population aged 20-55 years. This age range was chosen to align with the all-cause death rates for persons presenting chronic pain available in the literature. | Statistics Canada[10] |
| ***Third month post new MSK pain*** | | | | | |
| Probability of MSK pain resolving after three months | 55.39 | 45.41-64.75 | Dirichlet | Considering that all transition probabilities for each state must add up to 100%, this probability was obtained by subtracting this state's other transition probabilities from 100%. | Not applicable |
| Probability of MSK pain lasting more than three months | 44.60 | 35.25-54.59 | | Proportion of study participants whose MSK pain persisted after three months. | Heneweer, Aufdemkampe et al.[11] |
| Probability of dying while presenting new MSK pain | 0.011 | 0-0.061 | | Probability of death from any cause in the general Canadian population aged 20-55 years. This age range was chosen to align with the all-cause death rates for persons presenting chronic pain available in the literature. | Statistics Canada[10] |

**Table S3** Parameters used to perform the cost-utility analysis – Markov model (continued)

| Parameter | Values | 95% CI | Distribution Probabilistic analyses | Rationale | Source |
|---|---|---|---|---|---|
| **Transition probabilities [a]** | | **%** | | | |
| ***Chronic MSK pain*** | | | | | |
| Probability of chronic MSK pain lasting more than a month | 88.31 | 81.33-93.87 | Dirichlet | Considering that all transition probabilities for each state must add up to 100%, this probability was obtained by subtracting this state's other transition probabilities from 100%. | Not applicable |
| Probability of having a chronic MSK pain flare up | 11.21 | 5.72-18.08 | | Approximately 51% of participants with chronic non-specific back pain reported having a flare-up within a 6-month period. | Suri, Saunders et al.[12] |
| Probability of no longer having chronic MSK pain | 0.46 | 0-2.39 | | After a four-year follow-up, only 21.5% of participants presenting chronic pain at baseline were pain free, representing an annual recovery rate of 5.4%. | Elliott, Smith et al.[13] |
| Probability of dying while having chronic MSK pain | 0.025 | 0-0.30 | | Probability of death from any cause in women with self-reported MSK pain aged 20-55 years (17-year follow-up). | Nitter and Forseth[14] |
| ***Flare up*** | | | | | |
| Probability of flare up resolving after a month | 99.39 | 97.12-99.99 | Dirichlet | Considering that all transition probabilities for each state must add up to 100%, this probability was obtained by subtracting this state's other transition probabilities from 100%. | Not applicable |
| Probability of flare up lasting more than a month | 0.59 | 0-2.80 | | Proportion of participants with chronic non-specific back pain reporting flare ups lasting more than four weeks over a six-month period. | Suri, Saunders et al.[12] |
| Probability of dying while having a chronic MSK pain flare up | 0.025 | 0-0.30 | | Probability of death from any cause in women with self-reported MSK pain aged 20-55 years (17-year follow-up). | Nitter and Forseth[14] |
| ***Post flare up chronic MSK pain*** | | | | | |
| Probability of post flare up chronic MSK pain lasting more than a month | 93.06 | 87.24-97.16 | Dirichlet | Considering that all transition probabilities for each state must add up to 100%, this probability was obtained by subtracting this state's other transition probabilities from 100%. | Not applicable |
| Probability of having another chronic MSK pain flare up | 6.46 | 2.53-12.01 | | One-third of the participants in this study who had already had at least one flare up mentioned having had at least one new in the last six months. | Suri, Saunders et al.[12] |
| Probability of no longer having post flare up chronic MSK pain | 0.46 | 0-2.39 | | After a four-year follow-up, only 21.5% of participants presenting chronic pain at baseline were pain free, representing an annual recovery rate of 5.4%. | Elliott, Smith et al.[13] |
| Probability of dying while having post flare up chronic MSK pain | 0.025 | 0-0.30 | | Probability of death from any cause in women with self-reported MSK pain aged 20-55 years (17-year follow-up). | Nitter and Forseth[14] |
| **Costs, per month** | | **$ [b]** | | | |
| ***Post initial MSK pain*** | | | | | |
| Canadian Public Payer | 239.00 | 193.91-284.45 | Continuous with +/- 20% as SD | Costs associated with the use of public health care in the general population | Thanh, Tanguay et al.[15] |
| Canadian Society | 1,075.60 | 871.43-1,279.72 | | Costs associated with productivity loss and public health care in the general population. Of note, only persons between the ages of 18 and 65 were included in the productivity loss calculation. | |

**Table S3** Parameters used to perform the cost-utility analysis – Markov model (continued)

| Parameter | Values | 95% CI | Distribution Probabilistic analyses | Rationale | Source |
|---|---|---|---|---|---|
| **Costs, per month** | | **$ [b]** | | | |
| ***First month post new MSK pain*** | | | | | |
| Canadian Public Payer | 721.05 | 197.96-4,113.92 | Gamma | Includes pain-related public healthcare costs and costs associated with all-cause healthcare use in the general population. Mean cost was derived from data collected one month after the initial ED visit during the longitudinal follow-up carried out as part of a previous RCT. | Gagnon, Guertin et al. [under review] |
| Canadian Society | 1,650.36 | 895.75-4,599.40 | | Includes pain-related societal healthcare costs, costs associated with all-cause healthcare use in the general population, and costs associated with all-cause productivity loss in the general population. Mean cost was derived from data collected one month after the initial ED visit during the longitudinal follow-up carried out as part of a previous RCT. | |
| ***Second month post new MSK pain*** | | | | | |
| Canadian Public Payer | 280.20 | 227.84-333.35 | Gamma | Includes pain-related public healthcare costs and costs associated with all-cause healthcare use in the general population. Mean cost was derived from data collected three months after the initial ED visit during the longitudinal follow-up carried out as part of a previous RCT. | Gagnon, Guertin et al. [under review] |
| Canadian Society | 1,177.67 | 906.17-1,550.28 | | Includes pain-related societal healthcare costs, costs associated with all-cause healthcare use in the general population, and costs associated with all-cause productivity loss in the general population. Mean cost was derived from data collected three months after the initial ED visit during the longitudinal follow-up carried out as part of a previous RCT. | |
| ***Third month post new MSK pain*** | | | | | |
| Canadian Public Payer | 280.20 | 227.84-333.35 | Gamma | Includes pain-related public healthcare costs and costs associated with all-cause healthcare use in the general population. Mean cost was derived from data collected three months after the initial ED visit during the longitudinal follow-up carried out as part of a previous RCT. | Gagnon, Guertin et al. [under review] |
| Canadian Society | 1,177.67 | 906.17-1,550.28 | | Includes pain-related societal healthcare costs, costs associated with all-cause healthcare use in the general population, and costs associated with all-cause productivity loss in the general population. Mean cost was derived from data collected three months after the initial ED visit during the longitudinal follow-up carried out as part of a previous RCT. | |
| ***Chronic MSK pain*** | | | | | |
| Canadian Public Payer | 301.41 | 252.28-351.02 | Continuous with +/- 20% as SD | Includes pain-related public healthcare costs and costs associated with all-cause healthcare use in the general population. Using Canadian administrative health data, the authors calculated the direct healthcare cost of MSK pain paid by the Ontario (Canadian province) healthcare system. | Power, Perruccio et al.[16] |

**Table S3** Parameters used to perform the cost-utility analysis – Markov model (continued)

| Parameter | Values | 95% CI | Distribution Probabilistic analyses | Rationale | Source |
|---|---|---|---|---|---|
| **Costs, per month** | | **$ [b]** | | | |
| ***Chronic MSK pain (continued)*** | | | | | |
| Canadian Society | 2,791.43 | 2,357.77-3,228.62 | | Includes pain-related societal healthcare costs, costs associated with all-cause healthcare use in the general population, costs associated with all-cause productivity loss in the general population, and pain-related productivity loss costs. In their study, the authors estimated the annual cost of productivity loss among persons presenting with chronic pain at $16,960/person. | Thanh, Tanguay et al.[15] |
| ***Flare up*** | | | | | |
| Canadian Public Payer | 342.02 | 196.14-1,210.29 | Gamma | Includes pain-related public healthcare costs and costs associated with all-cause healthcare use in the general population. As part of their study, the authors calculated the monthly public healthcare cost of a person with chronic pain on a pain clinic waitlist. | Guerriere, Choinière et al.[17] |
| Canadian Society | 5,461.95 | 1,144.20-18,163.86 | | Includes pain-related societal healthcare costs, costs associated with all-cause healthcare use in the general population, costs associated with all-cause productivity loss in the general population, and pain-related societal costs. As part of their study, the authors calculated the monthly societal cost of a person with chronic pain on a pain clinic waitlist. Societal costs included were out-of-pocket, insurance, patient time lost, and family caregiver time lost. | |
| ***Post flare up chronic MSK pain*** | | | | | |
| Canadian Public Payer | 342.02 | 196.14-1,210.29 | Gamma | Includes pain-related public healthcare costs and costs associated with all-cause healthcare use in the general population. As part of their study, the authors calculated the monthly public healthcare cost of a person with chronic pain on a pain clinic waitlist. | Guerriere, Choinière et al.[17] |
| Canadian Society | 5,461.95 | 1,144.20-18,163.86 | | Includes pain-related societal healthcare costs, costs associated with all-cause healthcare use in the general population, costs associated with all-cause productivity loss in the general population, and pain-related societal costs. As part of their study, the authors calculated the monthly societal cost of a person with chronic pain on a pain clinic waitlist. Societal costs included were out-of-pocket, insurance, patient time lost, and family caregiver time lost. | |
| **Effectiveness** | **Utility score** | **95% CI** | | | |
| Post initial MSK pain | 0.824 | 0.504-0.989 | Beta | Mean utility score in the general population (Quebec, Canada). | Poder, Carrier, Kouakou[18] |
| First month post new MSK pain | 0.687 | 0.399-0.914 | | Mean utility score was derived from data collected one month after the initial ED visit during the longitudinal follow-up carried out as part of a previous RCT. | Gagnon, Guertin et al. [under review] |
| Second month post new MSK pain | 0.712 | 0.506-0.884 | | Mean utility score was derived from data collected three months after the initial ED visit during the longitudinal follow-up carried out as part of a previous RCT. | Gagnon, Guertin et al. [under review] |

**Table S3** Parameters used to perform the cost-utility analysis – Markov model (continued)

| Parameter | Values | 95% CI | Distribution Probabilistic analyses | Rationale | Source |
|---|---|---|---|---|---|
| **Effectiveness** | **Utility score** | **95% CI** | | | |
| Third month post new MSK pain | 0.712 | 0.506-0.884 | Beta | Mean utility score was derived from data collected three months after the initial ED visit during the longitudinal follow-up carried out as part of a previous RCT. | Gagnon, Guertin et al. [under review] |
| Chronic MSK pain | 0.579 | 0.057-0.986 | | Mean utility score measured in a sample of persons with chronic low back pain | Poder, Wang, Carrier[19] |
| Flare up | 0.479 | 0.016-0.978 | | Utility score measured in a sample of persons with chronic low back pain. This mean utility score corresponds to that of persons whose mean pain level over the last two weeks was between 7 and 10/10. This mean utility score was chosen to reflect the fact that, during a flare up, the pain level over the last two weeks is expected to be higher than usual. | Poder, Wang, Carrier[19] |
| Post flare up chronic musculoskeletal pain | 0.539 | 0.024-0.990 | | Utility score measured in a sample of persons with chronic low back pain. This mean utility score corresponds to that of persons whose worst pain level over the last two weeks was 7 to 10/10. This utility score was chosen to reflect the fact that, following an exacerbation, the pain level over the last two weeks is expected to have been higher than usual. | Poder, Wang, Carrier[19] |

CI: confidence interval; %: percentage; MSK: musculoskeletal; $: dollars; SD: standard deviation; ED: emergency department; RCT: randomized clinical trial

[a] All transition probabilities are per month

[b] All monthly costs are in 2024 Canadian dollars

**Table S4** Cost types included in each perspective

| Perspective | Cost types included |
|---|---|
| Canadian Public Payer [a] | - Emergency department visits<br>- Emergency department new visits for the same condition<br>- Ambulance transportation<br>- Hospitalizations<br>- Day surgeries<br><br>Each of the following cost type if covered by the Public Payer:<br><br>- Medical consultations (family doctors and specialists)<br>- Health professionals' consultations<br>- Imaging<br>- Prescription drugs<br>- Walking aids/Orthoses |
| Canadian Society [a] | - All costs included in the Canadian Public Payer perspective<br><br>All the following cost types, whether paid via insurers, out-of-pocket, a Workers' Compensation Program or a Public automobile insurance plan:<br><br>- All medical consultations, health professionals' consultations, imaging tests, prescription drugs and walking aids/orthoses made or purchased within the Private health system<br>- Over-the-counter drugs<br>- Other practitioners (e.g., massage therapist, osteopath)<br><br>Costs related to lost productivity:<br><br>- Lost productivity<br>- Patient time lost<br>- Family caregiver time lost |

[a] All cost types presented were included if available from the randomized clinical trial data and/or scientific literature

**Table S5** Uncertainty evaluation

| Type of uncertainty | Methods used to account for uncertainty |
|---|---|
| Model uncertainty | The Markov model was created by healthcare professionals with a good knowledge of the clinical evolution of the condition of interest (musculoskeletal pain). It was also validated by a panel of clinical experts who were not involved in its creation during a formal elicitation process (Table S1). |
| Parameter uncertainty | This type of uncertainty was addressed by performing a probabilistic sensitivity analysis via a Monte Carlo simulation. Each parameter (cost, efficiency, transition probabilities) was represented by a distribution of possible values, from which a value was randomly sampled during each iteration (n = 10,000 iterations, see Tables S2 and S3 for more information on the parameters and distributions used). |
| Uncertainty of results | Mean costs and utility scores used in the hybrid model were the same for both care models (i.e., did not favor one model over the other). It was therefore assumed that the only parameter that could influence the cost-utility of the two care models was the proportion of participants entering the "Post MSK pain" and "Chronic MSK pain" states at cycle 0 of the Markov model. Thus, four further probabilistic sensitivity analysis scenarios were carried out to ensure the robustness of the results obtained. Within these scenarios, only the transition probabilities of persons managed by a physiotherapist and an emergency physician were modified in a worst-case scenario type analysis. The assumptions tested in each scenario were as follows:<br>1. the probability of having musculoskeletal pain at the 3-month follow-up when presenting pain at the 1-month follow-up was 100%;<br>2. the probability of having musculoskeletal pain at the 1-month follow-up was doubled;<br>3. the probability of having musculoskeletal pain at the 1-month follow-up AND of having musculoskeletal pain at the 3-month follow-up when presenting pain at the 1-month follow-up were doubled;<br>4. the probability of having musculoskeletal pain at the 1-month follow-up was doubled AND the probability of having musculoskeletal pain at the 3-month follow-up when presenting pain at the 1-month follow-up was 100%. |

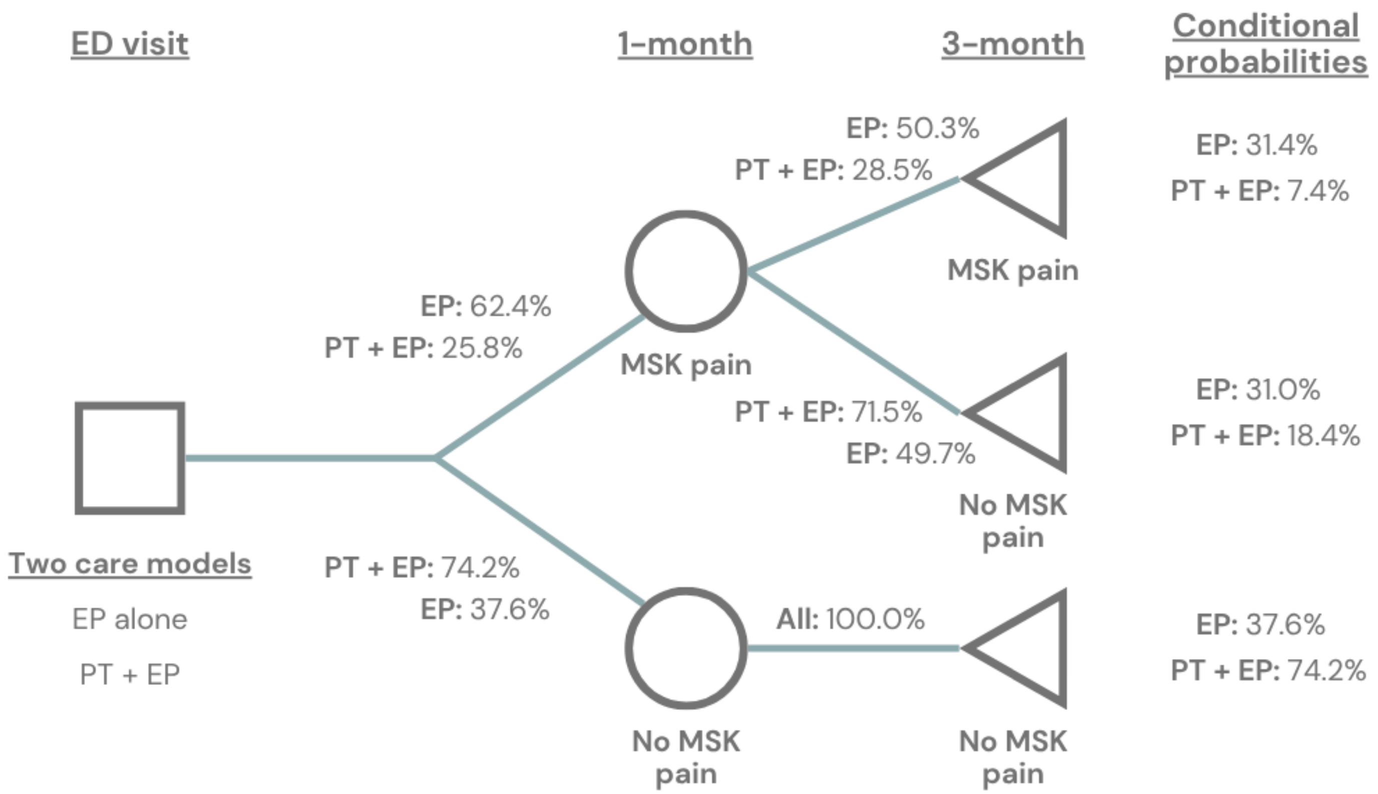


**Figure S1** Decision tree used as part of the hybrid mathematical model (with transition probabilities)

ED: emergency department; EP: emergency physician; PT: physiotherapist; MSK: musculoskeletal

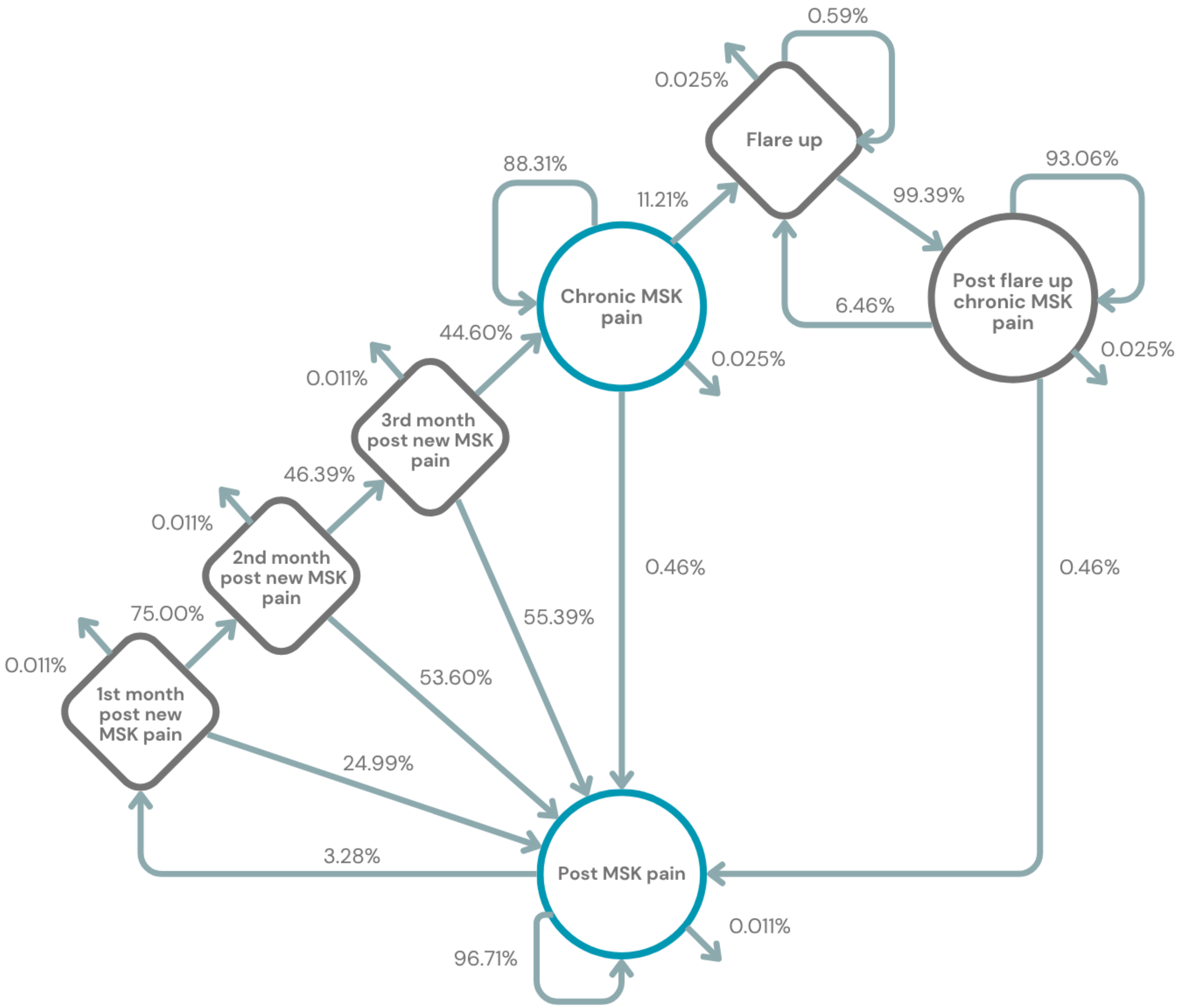


**Figure S2** Markov model used as part of the hybrid mathematical model (with transition probabilities)

MSK: musculoskeletal; 1st: first; 2nd: second; 3rd: third

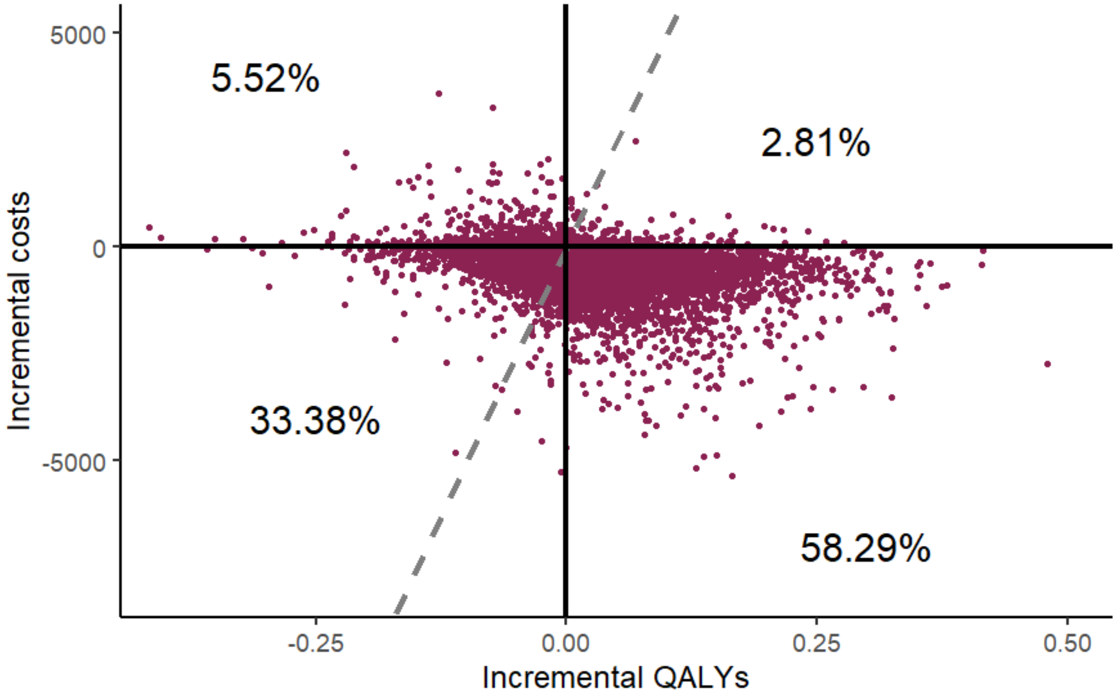


A.

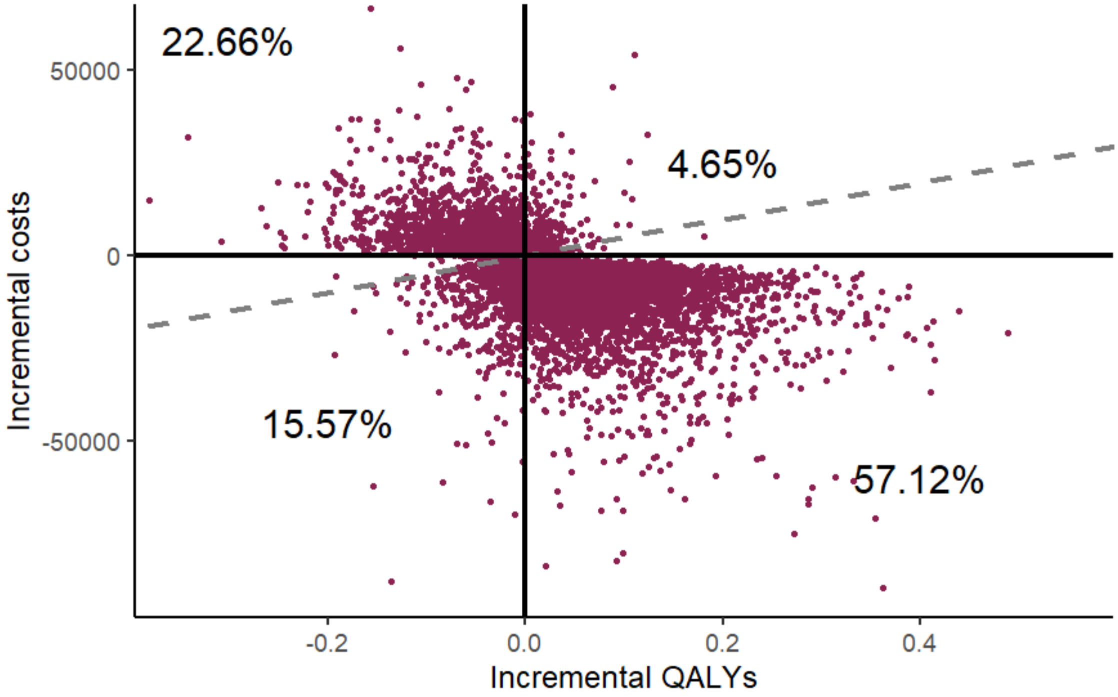


B.

**Figure S3** Two-year cost-effectiveness plane for each perspective – First assumption

<u>Assumption tested</u>: Probability of having musculoskeletal pain at the 3-month follow-up when presenting pain at the 1-month follow-up was 100% for all persons managed via the physiotherapist and emergency physician care model

A. Public Payer perspective
B. Societal perspective

QALYs: quality-adjusted life years

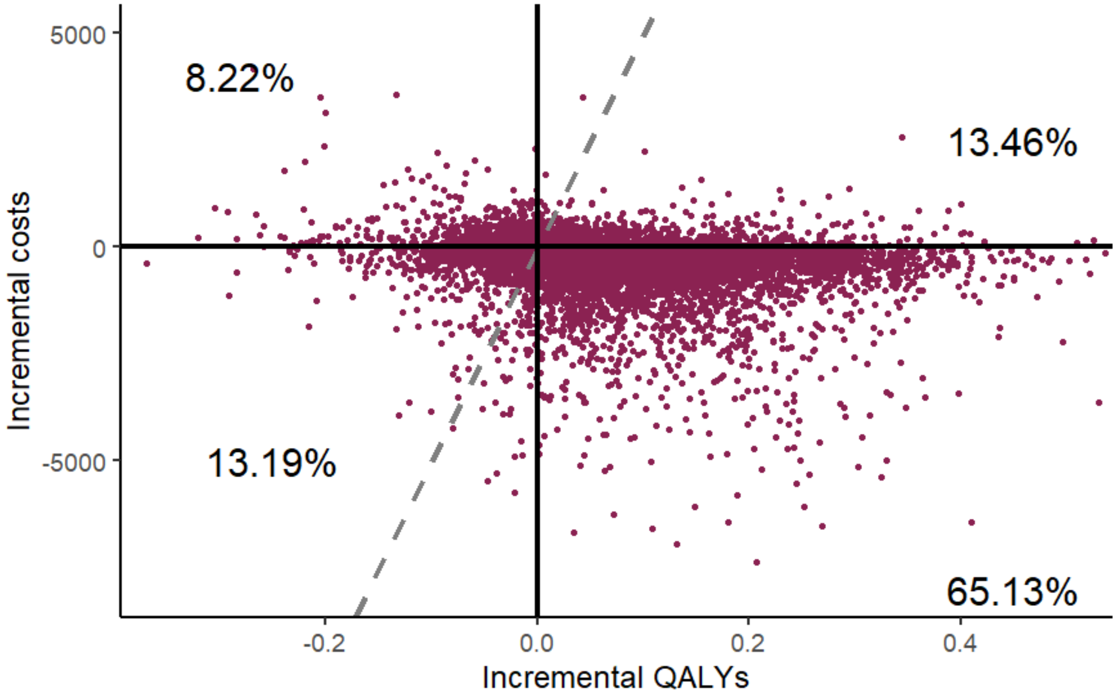


**A.**

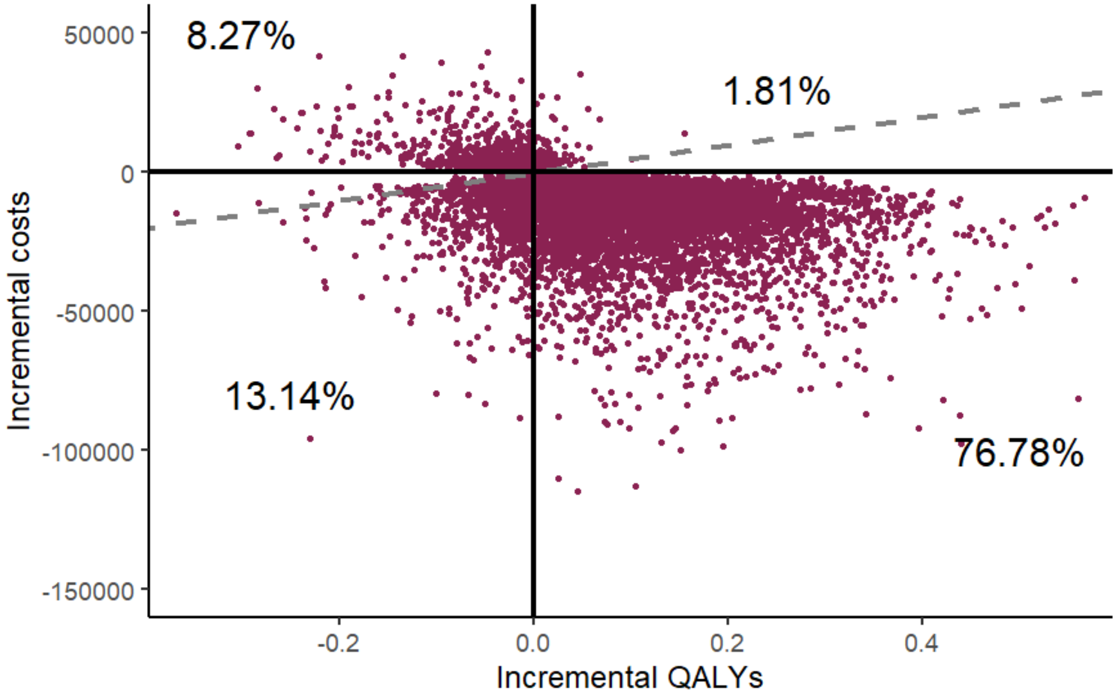


**B.**

**Figure S4** Two-year cost-effectiveness plane for each perspective – Second assumption

<u>Assumption tested</u>: Probability of having musculoskeletal pain at the 1-month follow-up was doubled for all persons managed via the physiotherapist and emergency physician care model

A. Public Payer perspective
B. Societal perspective

QALYs: quality-adjusted life years

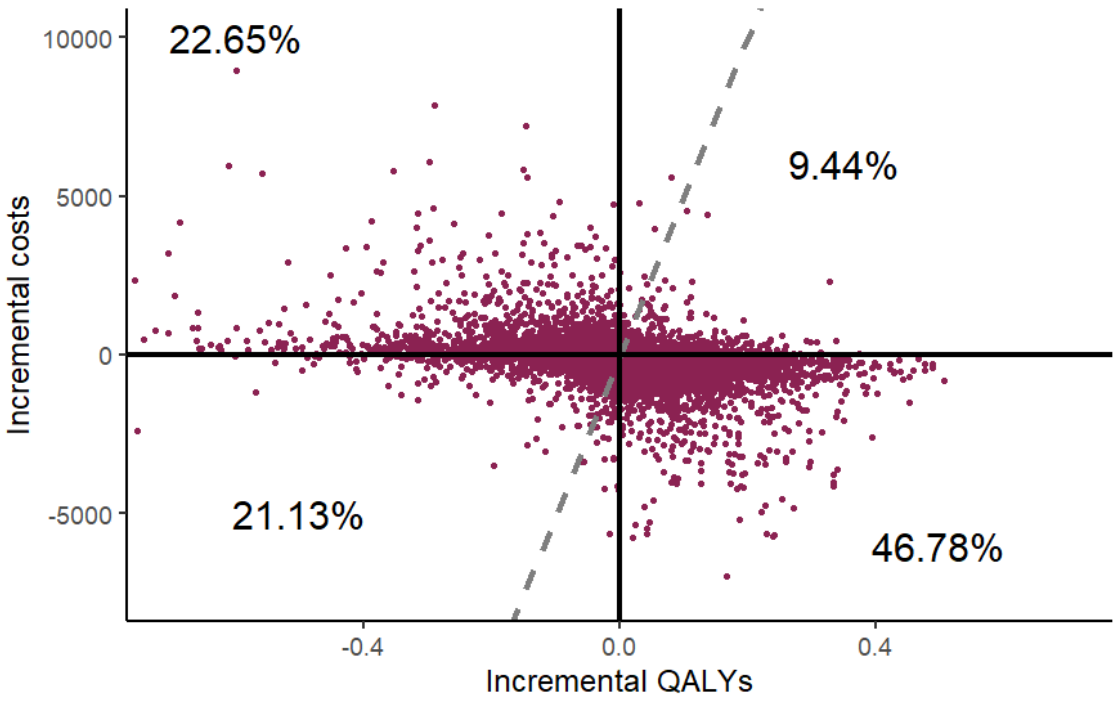


**A.**

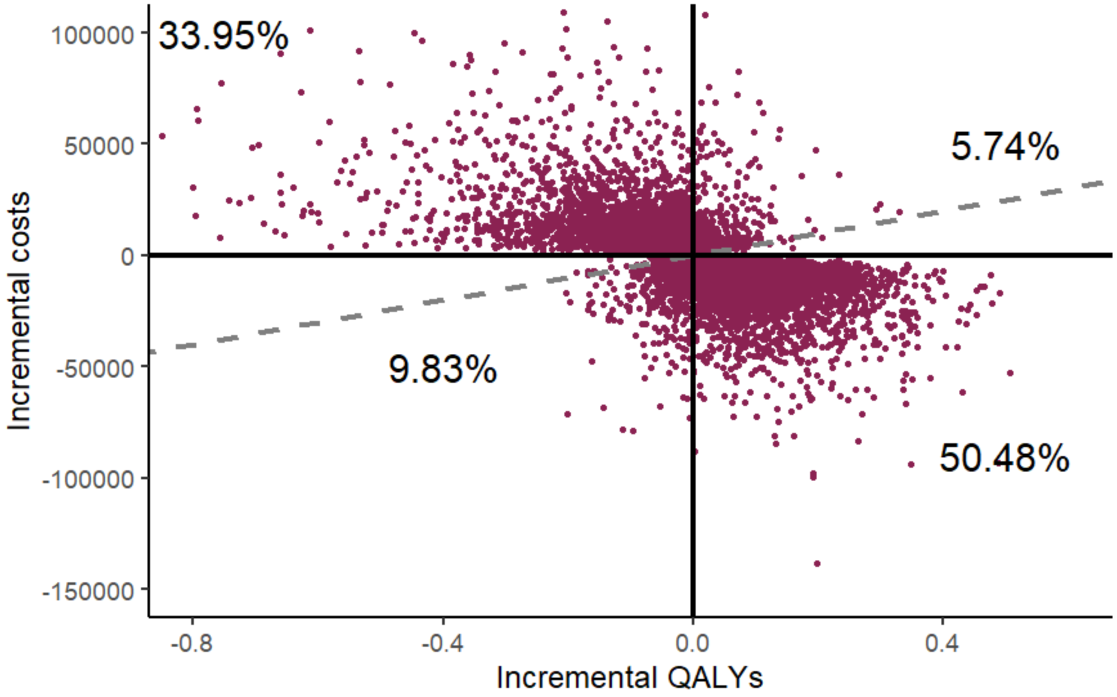


**B.**

**Figure S5** Two-year cost-effectiveness plane for each perspective – Third assumption

Assumption tested: Probability of having musculoskeletal pain at the 1-month follow-up AND of having musculoskeletal pain at the 3-month follow-up when presenting pain at the 1-month follow-up were doubled for all persons managed via the physiotherapist and emergency physician care model

A. Public Payer perspective

**B.** Societal perspective

QALYs: quality-adjusted life years

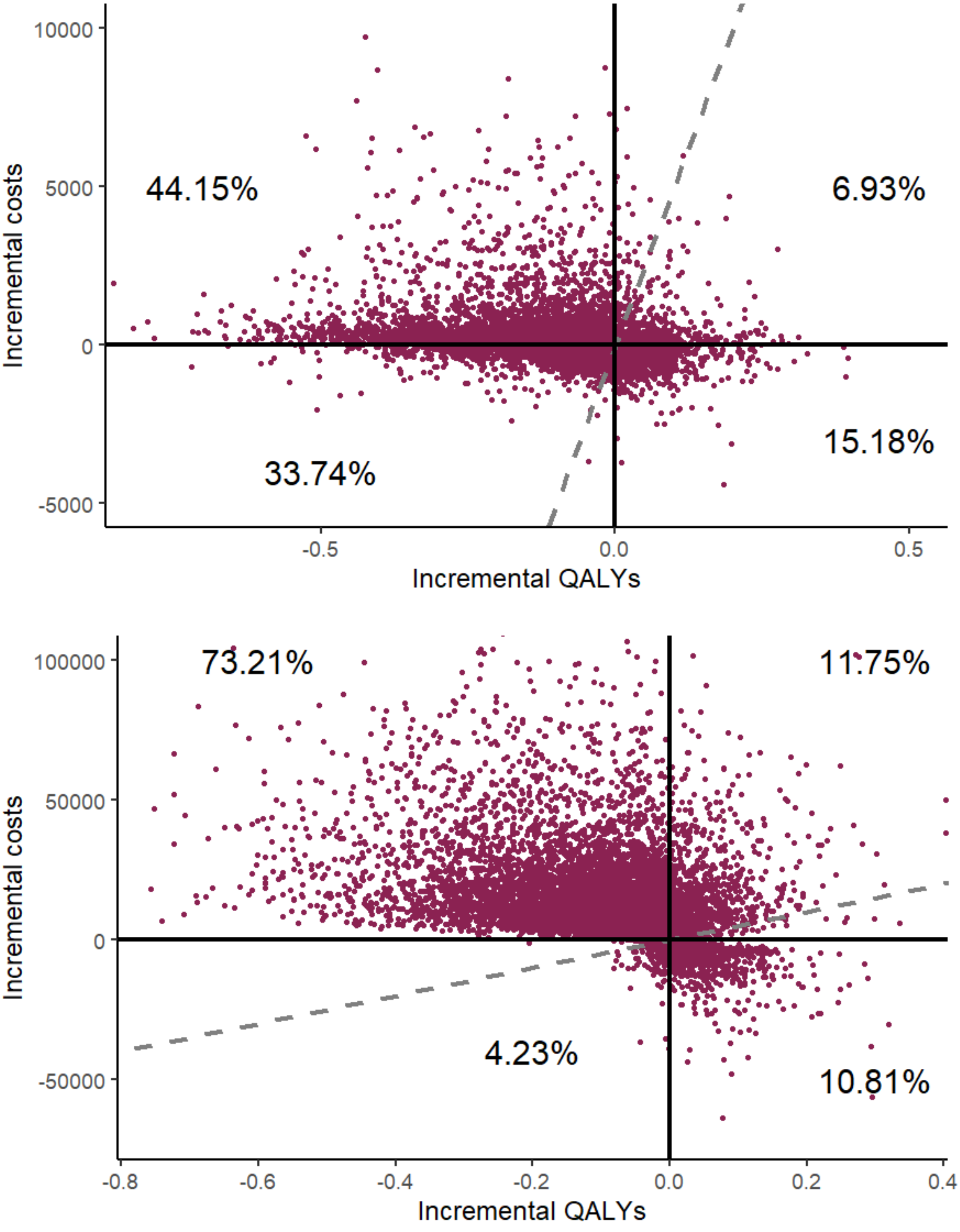


**Figure S6** Two-year cost-effectiveness plane for each perspective – Fourth assumption

<u>Assumption tested</u>: Probability of having musculoskeletal pain at the 1-month follow-up was doubled AND the probability of having musculoskeletal pain at the 3-month follow-up when presenting pain at the 1-month follow-up was 100% for all persons managed via the physiotherapist and emergency physician care model

A. Public Payer perspective
B. Societal perspective

QALYs: quality-adjusted life years
